# Medical students’ crisis-induced stress and the association with social support

**DOI:** 10.1101/2022.03.18.22272605

**Authors:** Vera M.A. Broks, Karen M. Stegers-Jager, Jeroen van der Waal, Walter W. Van den Broek, Andrea M. Woltman

**Affiliations:** Institute of Medical Education Research Rotterdam, Erasmus MC University Medical Centre Rotterdam, the Netherlands; Department of Public Administration and Sociology, Erasmus University Rotterdam, Rotterdam, the Netherlands

**Keywords:** medical students, COVID-19, crisis, stress, social support

## Abstract

**Background:** Medical schools are challenged to guard student wellbeing due to the potential negative impact of the COVID-19 outbreak on top of the already high prevalence of mental distress. Whereas social support is generally associated with less crisis-induced stress, it is unknown whether this applies to medical students during the COVID-19 outbreak.

**Objectives:** The impact of the COVID-19 outbreak on perceived stress of medical students was assessed by comparing their perceived stress levels during the outbreak to both their own baseline and the previous cohort’s pre-COVID-19 stress levels. Then, the association between social support and COVID-19 induced stress was assessed.

**Methods:** Dutch Year-1 medical students of cohort 2019 (*n*=99) completed the 14-item Perceived Stress Scale (PSS-14) at two time points: baseline (pre-COVID-19) and final measurement (COVID-19). Social support - emotional-informational support and club membership - was assessed during the final measurement. PSS and social support scores were compared to similar measurements of cohort 2018 (*n*=196). Students’ baseline stress levels, gender and study performance were controlled for when comparing two cohorts.

**Results:** Stress levels did not differ statistically significant between both pre-COVID-19 measurements of cohort 2018 and baseline cohort 2019. During the COVID-19 outbreak, cohort 2019 showed significantly higher stress levels compared to baseline (paired t-test: t=6.07, *p*<.001) and compared to cohort 2018 (linear regression: B=4.186, *p*<.001). Only during the COVID-19 outbreak, higher levels of social support - i.e. emotional-informational support (B=-0.75, *p*<.001) and club membership (B=-3.68, *p*<.01) - were associated with lower levels of stress.

**Conclusions:** During the COVID-19 outbreak, the perceived stress of medical students was higher - especially for students with lower levels of social support. Our results suggest that medical schools should optimize social support to minimize crisis-induced stress.

## Introduction

The prevalence of mental distress, i.e. anxiety-, depression-, or burnout symptoms, in medical students is high compared to their age-matched peers [1-3]. Approximately a quarter to one-third of medical students shows symptoms of depression [4, 5], and roughly 40% shows burnout symptoms [6]. These mental problems can be caused by stress [7]. A recent stressor in the shape of a crisis, is the COVID-19 outbreak. The outbreak’s potential negative impact on mental wellbeing combined with the already high prevalence of mental problems in medical students, exacerbates the challenge medical schools face to guard their students’ wellbeing [8]. Research regarding factors related to higher stress levels during a crisis - or in other situations in which stressors increase - will enable medical schools to limit the negative impact of such crises on student wellbeing. Social support is possibly one of the factors that is associated with crisis induced stress [9-11]. Therefore, the present study has two objectives. The first objective is to investigate whether the COVID-19 outbreak impacted perceived stress of medical students. The second objective is to investigate the association between social support and COVID-19 induced stress for medical students.

The COVID-19 outbreak has disrupted everyday life, which negatively impacted the mental wellbeing of the general population [12-14]. Compared to the general population, especially students reported mental health problems during the outbreak [15]. A possible explanation is that student life and its social aspects were affected by measurements regarding social distancing, lockdown and if necessary quarantine [16], including the transition to online education [17]. However, for medical students, studies show mixed results regarding the impact of the COVID-19 outbreak on wellbeing. A systematic review reports that anxiety levels in medical students did not increase during the outbreak [18]. Whereas other studies report higher levels of burnout symptoms and stress for medical students during the outbreak [19], especially female students [20, 21]. However, these studies often consisted of self-reported increased stress levels without a baseline measurement [20, 21], or did not correct for probable changes of stress levels throughout the academic year regardless of the outbreak [19]. Finally, the role of academic performance was not taken into account in previous studies while a negative relationship between study performance and stress is known for medical students [22]. In summary, previous studies with respect to the impact of the COVID-19 outbreak often lack controls for baseline measurements and student performance.

According to the stress-buffering model, the negative impact of a stressful event on wellbeing is stronger for individuals with less social support [10, 11]. Firstly, because social support can prevent that someone appraises an event as highly stressful. Secondly, because social support alleviates the impact of the appraised stress by, for example, offering a solution or reducing the importance. Social support refers to people who someone can turn to in times of crisis – such as family, friends, colleagues or neighbours. A literature review shows that after a hurricane, tsunami or terrorist attack, social support is associated with more resilience [23]. During the COVID-19 outbreak, literature with respect to the general population indicates that social support is associated with a decreased sense of loneliness [24], and more resilience [25]. For adolescents and college students, social support in times of COVID-19 is associated with less depression and anxiety symptoms [26], and less stress symptoms [27]. For medical students, social support has been shown to be positively linked to mental wellbeing [28-34]. However, to the best of our knowledge, it is not yet known whether social support is associated with crisis-induced stress among medical students, especially in a crisis that strains social contacts by all kinds of social distancing measures.

The COVID-19 outbreak offered the unique opportunity to assess crisis-induced stress on a large scale for medical students. Existing studies cannot provide solid evidence of shifted stress levels during the COVID-19 outbreak due to lack of required baseline measurements and controls for study performance. Even though medical schools are unable to resolve the present COVID-19 crisis or any future crisis, they might be able to help limit the negative consequences on – the already relatively low - student wellbeing. In order to get more insights in how medical schools can provide the right support to their students in times of crisis, we examined the following research questions:

1. What is the impact of the COVID-19 outbreak on medical students’ perceived stress compared to both their own baseline stress level and compared to the stress level of the previous cohort, while controlling for gender and study performance?
2. Does social support moderate the effect of the COVID-19 outbreak on the perceived stress of medical students?

## Methods

### Context

The present study was conducted with Year-1 Bachelor students of Erasmus MC Medical School in the Netherlands. Dutch medical schools consist of a 3-year Bachelor’s- and 3-year Master’s program. At Erasmus MC Medical School, the Bachelor curriculum is composed of preclinical training in thematic blocks and competence-based learning lines. Each year, 6o credits under the European Credit Transfer System (ECTS) can be obtained, resulting in 180 credits for the complete Bachelor program. Grades are based on a 10-point scale from 1 to 10 (maximum) where 5.5 is the minimum to pass. In March 2020, the COVID-19 outbreak started to impact everyday life in the Netherlands. For the Bachelor students of Erasmus MC Medical School, this entailed that all classes were dismissed and online education became the new standard.

### Participants and procedure

All Year-1 Bachelor students from Erasmus MC Medical School, who enrolled in cohorts 2018 (409 students) and 2019 (408 students) were invited to participate by completing a questionnaire regarding perceived stress in December/January (baseline measurement – online questionnaire) and May (final measurement – cohort 2018 on paper and online possible, cohort 2019 online) of their first academic year. During the final measurement, social support was measured beside perceived stress. The sample in the present study consisted of students who completed both questionnaires. Only the final measurement of cohort 2019 took place during the COVID-19 outbreak (see Fig 1A). Students provided informed consent for the data collected by questionnaires. They also agreed to link questionnaire results to relevant data from the student administration. The university student administration provided data regarding gender and study performance up to the final measurement. Since these data only were reported on an aggregated level for the complete cohorts, no individual consent was required. The study was carried out in accordance with the Declaration of Helsinki and was deemed exempt from review after evaluation by the Medical Ethics Committee of Erasmus MC Rotterdam (MEC-2019-0448).

**Fig 1.**
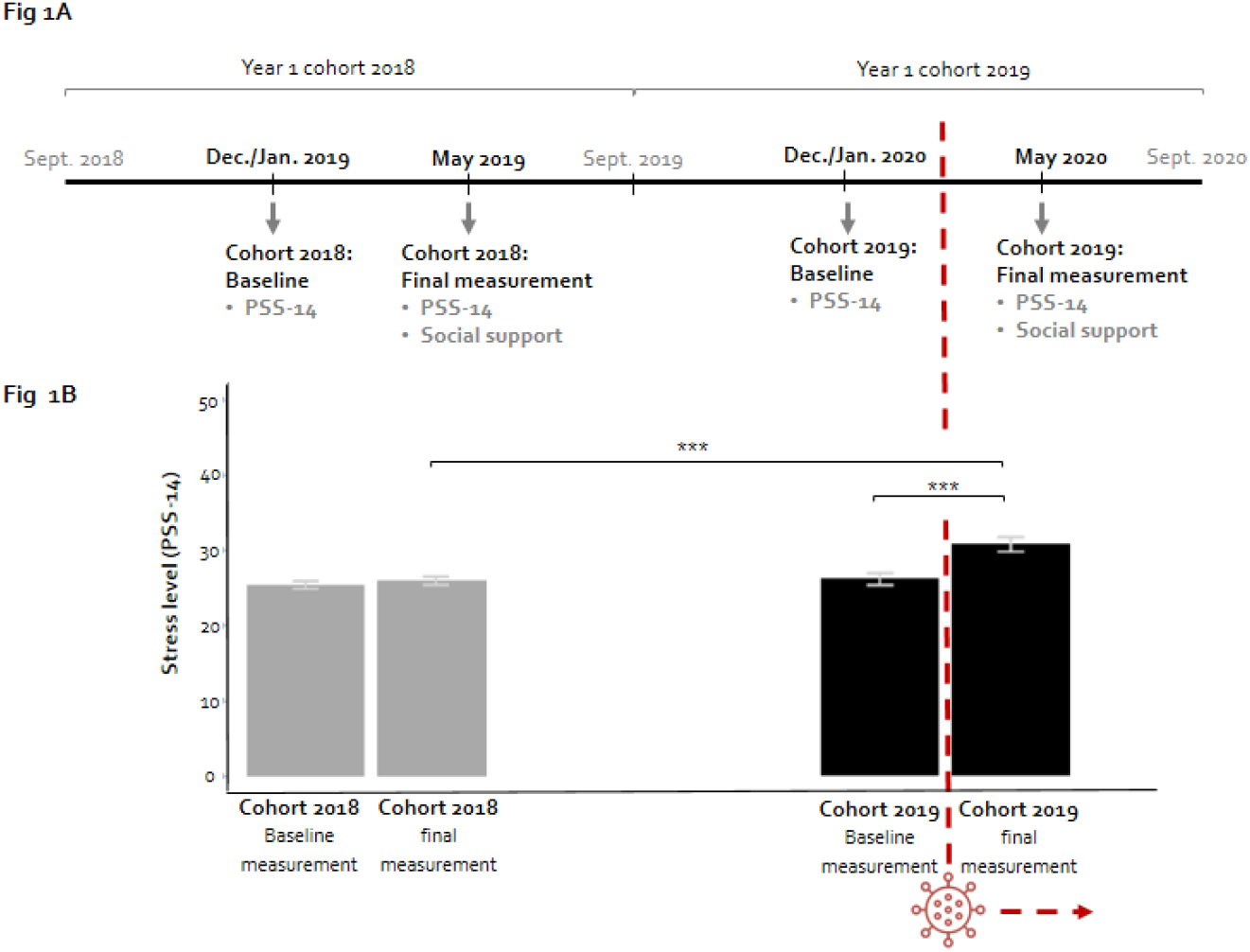
Timeline of data collection and mean perceived stress level for each measurement. **Fig 1A. Timeline of data collection**. The COVID-19 pandemic started to impact everyday life and medical school during Year-1 of cohort 2019 (March 2020). **Fig 1B. Mean perceived stress levels including error bars**. Mean stress-levels are shown for the baseline measurement (mean=25.38, SE=0.51) and the final measurement (mean=26.00, SE=0.54) for cohort 2018 and baseline measurement (mean=26.05, SE=0.79) and the final measurement (mean=30.65, SE=0.99) for cohort 2019. Differences are assessed within a cohort with a paired t-test. The difference between the final measurement of cohort 2018 and cohort 2019 is assessed in a linear regression model while controlling for baseline measurement, gender and study performance (Table 2 – Model 1). Significant differences between measurements are shown: *** p<.001.

### Measurements

#### Student characteristics

Student characteristics taken into account in the present study are gender and study performance. Gender is categorized as male or female. Student performance is operationalized by measuring whether students obtained all possible credits up until the final measurement in May of Year-1 (yes/no). The maximum number of obtainable credits up until the final measurement was equal to 38 credits for cohort 2018 and equal to 25 credits for cohort 2019. This difference in credits was due to the fact that for cohort 2019, some exams were postponed due to the COVID-19 outbreak along with the transition to online education.

#### Perceived stress (PSS-14)

Perceived stress was measured with a Dutch version of the 14-item perceived stress scale (PSS-14) [35, 36]. As recently described, this questionnaire is appropriate to measure stress responses [37], and measures general distress and someone’s ability to cope with this stress. The questionnaire focuses on feelings during the last month, therefore, every item starts with “In the last month…”, for example “In the last month, how often have you been upset because of something that happened unexpectedly?”. The items are scored on a 5-point Likert scale ranging from 0 (never) to 4 (very often). The maximum score on the PSS-14 is 56.

#### Social support

Social support was assessed with questions regarding emotional-informational support and club membership to cover different types of social support that are relevant to students in the Netherlands [38]. To measure emotional-informational support, four items of the MOS social support scale were selected from the subscale emotional-informational support [39] (S1 Table). For the selection of the items, the following was taken into account: an equal distribution of both emotional and informational items, item-scale correlations and applicability to our sample of medical students. The selected items focus on the availability of different forms of social support: for example, having someone to listen to you, to offer you advice or to share your thoughts with. The items are scored on a 5-point Likert scale ranging from 0 (never) to 4 (always). The maximum score on emotional-informational support is 16. Alpha reliability measures were computed in order to assess internal consistency for this adjusted scale (S1 Table). The second operationalization of social support was club membership. Club membership was measured with the closed yes/no question “Are you member of a hobby club, sports club or a leisure club?”.

### Statistical analysis

Analyses were performed on results of students that completed both the baseline questionnaire and final questionnaire during Year-1. First, the samples of the two included cohorts were compared regarding response rates, gender, study performance, social support and stress levels. T-tests and Wilcoxon rank-sum tests were used for the comparison on normally and non-normally distributed continuous variables respectively. Dichotomous variables were compared between cohorts with chi-square tests. Then, to study the impact of COVID-19 on perceived stress, paired t-tests were performed on baseline stress levels and related final measurement stress levels of both cohort 2018 and cohort 2019. In addition, stress levels during the final measurement of cohort 2018 (pre-COVID-19) and cohort 2019 (COVID-19) were compared in a linear regression model while controlling for baseline stress level, gender and study performance. The effect of social support on perceived stress was assessed by adding emotional-informational support and club membership to the linear regression model. Finally, the interaction terms of social support and cohort were added to the linear regression model to investigate whether the relation between social support and perceived stress was different during the COVID-19 outbreak. Post-hoc linear regression analysis was performed for both cohorts to study whether emotional-informational support and club membership were complementary to each other in relation to stress levels. To this end, groups based on social support were formed by crossing the presence of club membership (yes/no) with emotional-informational support (high/low). Emotional-informational support was considered low when students’ scores were in the 25^th^ percentile of the complete sample.

## Results

### Cohort characteristics

The response rate was lower in cohort 2019 compared to cohort 2018 (24% vs. 48%, Table 1). Proportions of female students and of students who acquired all obtainable credits until the final measurement were comparable between cohorts (see Table 1). Though emotional-informational support scores remained the same (Table 1), a chi-square test showed that the percentage of students that was member of a club was significantly lower in cohort 2019: 56% in cohort 2019 compared to 68% in cohort 2018 (*χ2*=4.150, df=1, *p*<0.05). The mean baseline stress level was comparable between cohorts, but the mean stress level during the final measurement was significantly higher for cohort 2019 than for cohort 2018 (*t*=4.134, df=158.69, p<.001).

**Table 1.** Descriptive statistics of variables included in the analyses.

|  | Cohort 2018 |  | Cohort 2019 |  | Comparison of cohorts (samples) |
| --- | --- | --- | --- | --- | --- |
|  | Total | Sample | Total | Sample |  |
| **Number of students (% of total)** | 409 (100) | 196 (48) | 408 (100) | 99 (24) | $\chi^2= 48.529$, $df=1$, $p<.001$ |
| **Female students: %** | 74 | 79 | 68 | 74 | $\chi^2= 0.787$, $df=1$, $p=0.375$ |
| **Students with all credits obtained until final measurement: %** | 66 | 71 | 65 | 67 | $\chi^2= 0.378$, $df=1$, $p=0.539$ |
| **<u>Social support</u>** |  |  |  |  |  |
| **Median emotional-informational support: median/total (range)<sup>a</sup>** | - | 14/16 (3-16) | - | 14/16 (2-16) | $W=10250$, $p=0.419$ |
| **Club member: % yes** | - | 68 | - | 56 | $\chi^2= 4.150$, $df=1$, $p<.05$ |
| **<u>Stress level (PSS-14)</u>** |  |  |  |  |  |
| **Mean baseline measurement** | - | 25.38 | - | 26.05 | $t=-0.710$, $df=181.62$, $p=0.479$ |
| **Mean final measurement** | - | 26.00 | - | 30.65 | $t=4.134$, $df=158.69$, $p<.001$ |

<sup>a</sup> Data are not normally distributed, non-parametric Wilcoxon rank-sum test is used to compare cohorts

### Perceived stress levels

A paired t-test showed that the perceived stress levels of cohort 2019 significantly increased from 26.05 to 30.65 (t=6.07, df=98, *p*<.001), whereas the stress levels of cohort 2018 did not significantly differ between the baseline and final measurement (Fig 1B). This indicates that the perceived stress levels of students significantly increased during the COVID-19 outbreak.

To assess the effect of the COVID-19 outbreak, we not only compared stress levels *within* cohorts, we also compared the stress levels during the final measurement *between* two cohorts of medical students. With respect to the control variables, a higher baseline stress level and not having obtained all obtainable credits were related to higher stress levels during the final measurement (Table 2 – Model 1). When controlling for baseline stress level, gender and study performance, a significantly higher perceived stress level was visible for cohort 2019 (COVID-19) compared to cohort 2018 (Pre-COVID-19; B_cohort_=4.186, 95%- CI: 2.608 – 5.764, *p*<.001, Table 2 – Model 1 and Fig 1B). Compared to cohort 2018, the perceived stress levels for students of cohort 2019 – during the COVID-19 outbreak – were on average approximately 4 units higher on the Perceived Stress Scale (ranging from 0 to 56).

**Table 2.** Results linear regression model with outcome variable stress level (PSS-14) in the final measurement.

|  | Model 1 |  | Model 2 |  | Model 3 |  |
| --- | --- | --- | --- | --- | --- | --- |
|  | B [95% CI] | sig. | B [95% CI] | sig. | B [95% CI] | sig. |
| *Intercept* | 11.353 [8.000 – 14.707] | - | 19.004 [14.022 – 23.985] | - | 15.370 [9.740 – 20.999] | - |
| PSS (first measurement) | 0.616 [0.511 – 0.721] | *** | 0.567 [0.461 – 0.672] | *** | 0.579 [0.474 – 0.684] | *** |
| Gender (female)<sup>a</sup> | 1.766 [-0.061 – 3.594] | . | 2.231 [0.433 – 4.030] | * | 2.163 [0.384 – 3.943] | * |
| All credits obtained<sup>b</sup> | -3.372 [-5.025 – -1.718] | *** | -3.637 [-5.260 – -2.016] | *** | -3.427 [-5.041 – -1.813] | *** |
| Cohort (2019; COVID-19)<sup>c</sup> | 4.186 [2.608 – 5.764] | *** | 3.894 [2.341 – 5.446] | *** | 12.442 [5.775 – 19.109] | *** |
| Emotional-informational (E-I) support |  |  | -0.417 [-0.659 – -0.177] | *** | -0.230 [-0.532 – 0.0714] |  |
| Club member (yes)<sup>d</sup> |  |  | -1.636 [-3.197 – -0.077] | * | -0.501 [-2.429 – 1.428] |  |
| Cohort (2019) * E-I support |  |  |  |  | -0.516 [-1.004 – -0.027] | * |
| Cohort (2019) * Club member (yes) |  |  |  |  | -3.180 [-6.333 – -0.028] | * |
| Adjusted R<sup>2</sup> | 0.436 |  | 0.462 |  | 0.474 |  |

Note: Dependent variable: stress level during the final measurement: score on the PSS-14 (min=0, max=56). “B” refers to the unstandardized regression coefficient together with the 95% confidence interval (95% CI). For each regression coefficient, the table shows whether it significantly deviates from 0 (H<sub>0</sub>: B=0, H<sub>A</sub>: B≠0) in the column “sig.” using the following codes: \*\*\* p<.001, \*\*p<.01, \*p<.05, . p<.1. <sup>a</sup>Reference category: male gender. <sup>b</sup>Reference category: *not all possible credits obtained*. <sup>c</sup>Reference category: *cohort 2018 (pre-COVID-19)*. <sup>d</sup>Reference category: *no club* *membership*.

### Social support

After including social support, the difference in perceived stress levels between cohorts remained present (B_cohort_=3.894, 95%-CI: 2.341 – 5.446, *p*<.001, Table 2 – Model 2). This means that when taking into account social support, perceived stress levels were still significantly higher during the COVID-19 outbreak. In addition, when a student experienced more emotional-informational support, perceived stress levels were lower (B_E-I support_=-0.417, 95%-CI: −0.659 – −0.177, *p*<.001, Table 2 – Model 2). Also, students who were member of a club had significant lower stress levels compared to their fellow students who were not member of a club (B_club member_=-1.636, 95%-CI: −3.197 – −0.077, *p*<.05, Table 2 – Model 2).

Finally, to investigate whether social support is associated with the effect of the COVID-19 outbreak on perceived stress levels, the moderating effects of emotional-informational support and club membership with cohort were included. Both the effects of emotional-informational support (B_E-I support*cohort_=-0.516, 95%-CI: −1.004 – −0.027, *p*<.05) and club membership (B_club member*cohort_=-3.180, 95%-CI: −6.333 – −0.028, *p*<.05; Table 2 – Model 3) were significantly different for cohort 2019 (COVID-19) compared to cohort 2018 (pre-COVID-19). For cohort 2019, a significant decrease in stress levels was visible as emotional-informational support increased (Fig 2A). For cohort 2018 a slight decrease in stress level was visible too, however, this decrease was not statistically significant. Regarding club membership, only cohort 2019 demonstrated a significant difference in stress level between students who were a club member or not (Fig 2B). The results indicate that for cohort 2019, emotional-informational support and club membership were complementary to each other in relation to stress levels since the effects exist beside each other. In line, post-hoc analysis showed that for cohort 2019, emotional-informational support and club membership were complementary to each other in relation to stress levels. Students with only one of the two types of social support – i.e. club membership *or* high emotional-informational support - showed significantly higher stress levels compared to students with both types of social support (Fig 3). For cohort 2018, differences in stress levels based on social support were not present.

**Fig 2.**
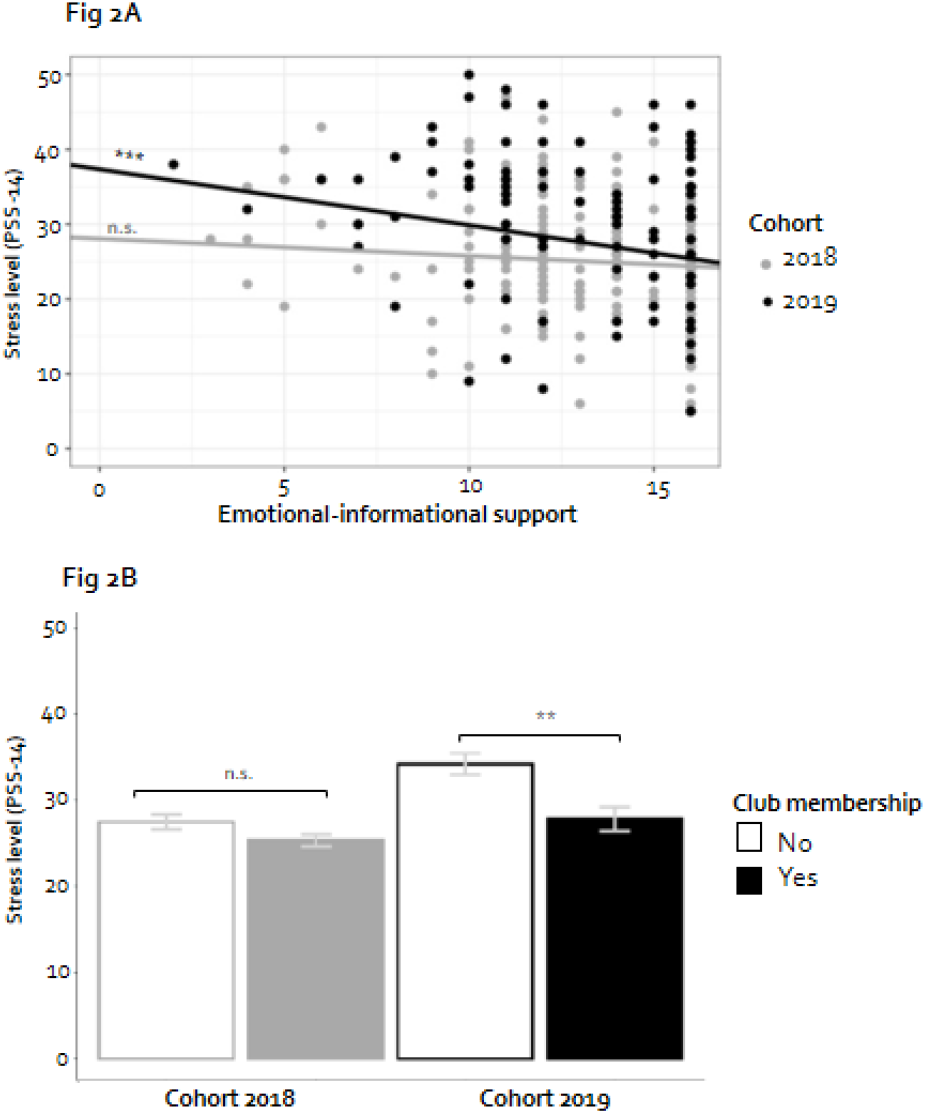
Social support and perceived stress level during the final measurement per cohort. **Fig 2A. Emotional-informational support**. Each dot represents an observation and demonstrates the perceived stress level and the level of emotional-informational support of an individual student. The slope of the regression line represents the effect of emotional-informational support on stress level. The slope of the regression line in grey for cohort 2018 is equal to −0.23 (non-significant, Table 2 – Model 3). The slope of the regression line in black for cohort 2019 is −0.75 (p<.001; p-value is based on a post-hoc analysis), which equals the sum of the regression coefficients for emotional-informational support and emotional-informational support*cohort 2019= −0.23-0.52=-0.75 (Table 2 – Model 3). **Fig 2B. Club membership**. Observed mean perceived stress levels including error bars (M±SE) for students without and with club membership. In the linear regression model, for cohort 2018, no significant effect is present for club membership (coefficient club member = −0.50, non-significant, Table 2 – Model 3). For cohort 2019, the linear regression model shows a difference of −3.68 (p<.01; p-value is based on a post-hoc analysis), which is the sum of the regression coefficients for club member and club member*cohort 2019: −0.50-3.18=-3.68 (Table 2 – Model 3).

**Fig 3.**
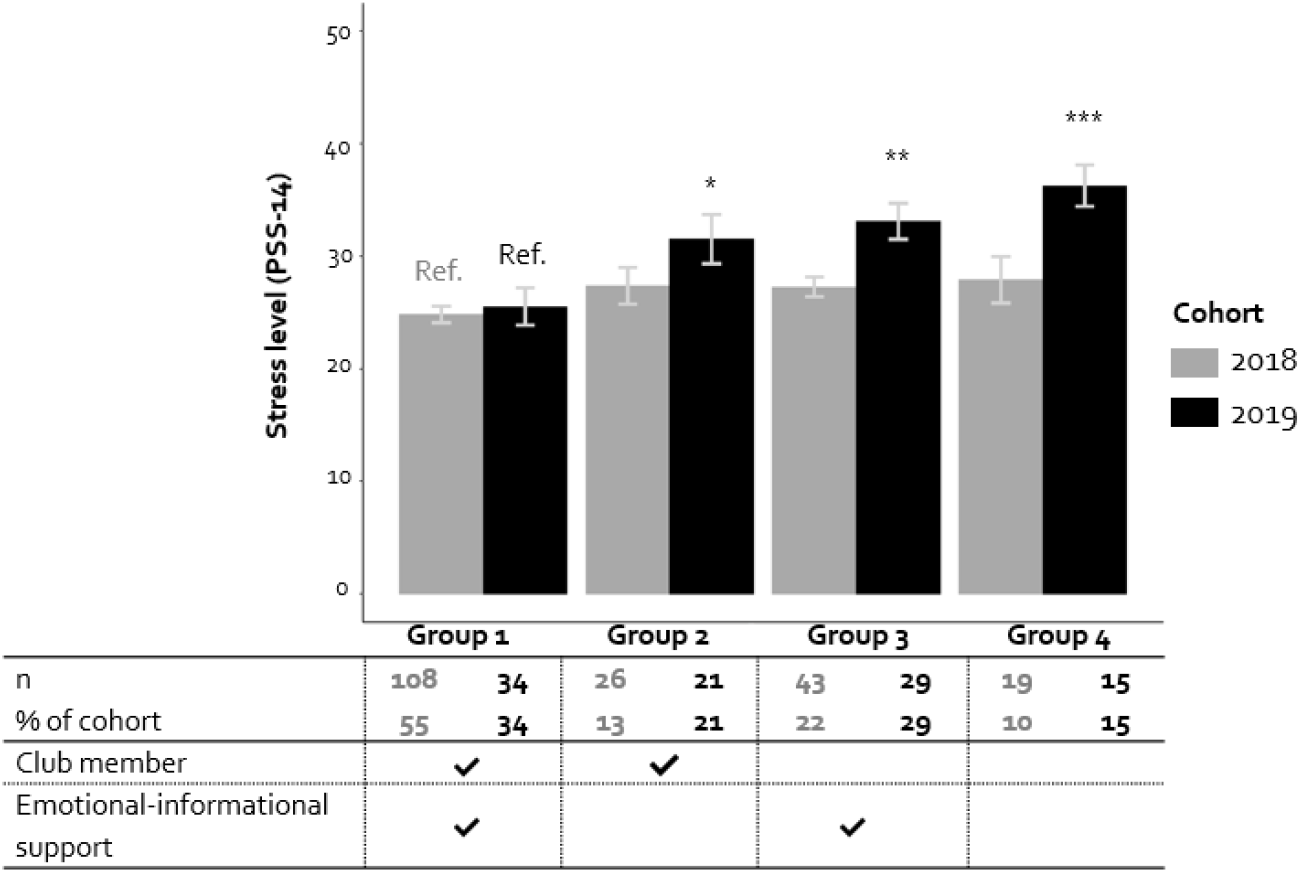
Perceived stress level of groups during the final measurement based on club membership and emotional-informational support. Mean perceived stress level for groups based on club membership and emotional-informational support for cohort 2018 and cohort 2019 including error bars. Emotional-informational support is unchecked if the score is ≤25th percentile of the complete sample (score ≤11). For each cohort separately, the stress level of groups 2 to 4 are compared to the stress level of group 1 in a post-hoc linear regression analysis (reference group=Ref.). For cohort 2018, group 2 to 4 do not differ significantly from group 1 regarding stress level. For cohort 2019, group 2 to 4 do differ significantly from group 1 regarding stress level: *p<.05, **p<.01, *** p<.001, respectively.

## Discussion

The present study demonstrates that the perceived stress levels of medical students were significantly higher during the COVID-19 outbreak – compared to their own pre-COVID-19 baseline stress levels as well as to stress levels of the previous cohort. During the outbreak, students experiencing less emotional-informational support or not being member of a club, showed higher perceived stress levels compared to their fellow students with more emotional-informational support or who were member of a club. The results indicate that during times of crisis, social support is associated with less perceived stress of medical students.

Our finding that stress levels of medical students during the COVID-19 outbreak were higher compared to pre-COVID-19 stress levels, corresponds with previous studies reporting higher stress levels during the outbreak [19-21]. We found that students’ stress levels increased compared to both their own baseline stress levels as well as compared to the stress levels of a previous cohort, including controls for gender and study performance. Also, we could take into account potential fluctuations during the academic year regardless of the pandemic, as reported in a previous study [40]. The mean stress level reported in the present study during the outbreak was higher than the mean stress level of Dutch medical students after increased performance standards [36]. Yet, the currently reported stress levels did not exceed stress levels of medical students in the US after the implementation of a new curriculum [41]. The reason why the COVID-19 outbreak elevated stress levels goes beyond the scope of the present study, but one might speculate about a mix of academic uncertainty [8, 17], online education [20], blurred study-home boundaries and social isolation [42]. Students’ stress levels are subjected to a variety of factors and thereby context-specific. This context-specificity of students’ stress levels illustrates the importance of baseline measurements to make valid comparisons. Such baseline measurements could also serve future research on the long-term effects of this COVID-19 induced stress.

In line with the stress-buffering model [10, 11], we found that only during the COVID-19 outbreak, higher levels of social support were associated with lower stress levels for medical students. This shows that also during a crisis in which social contacts are strained by social distancing measures, social support is associated with lower stress levels. This mechanism is also corroborated by a recent study in which medical students report that their own most effective strategies to lower the negative impact of stress include de-stress through friends and family [43]. Studies report that in times of the COVID-19 outbreak, social support is positively associated to wellbeing in adolescents [26], college students[27], and the general population [24]. These present study adds to these findings in two ways. First, by providing evidence that this also applies to medical students; a group that already experiences higher levels of mental distress compared to their peers [1-3]. Second, by showing that the association between social support and stress level also applies in times of crisis where social support is strained due to social distancing measurements [12, 13]. Even though the present study did not reveal lower levels of emotional-information support during the COVID-19 outbreak, the number of students who reported being member of a club did decrease from 68% to 56%. This decrease is possibly the result of measures taken to limit the spread of the coronavirus, thereby making it impossible for clubs to gather with their members. A potential explanation why emotional-informational support did not decrease is that students moved back home to their parents’ house where emotional-informational support was still available to them since close family is a source of social support for Dutch students [38].

The present study illustrates how two forms of social support – emotional-informational support and club membership - are complementary to each other in relation to reported stress levels during the COVID-19 outbreak. A possible explanation for this finding can be found in a theoretical model for mechanisms linking social support to health, where primary and secondary social resources are distinguished, described as *intimates* and *knowledgeable others* respectively [44]. According to this model, the two sources of social support each have their own attributions to buffering the impact of stressors. *Intimates* buffer the stress by companionate presence, offering care and instrumental assistance whereas *knowledgeable others* buffer stress by enabling ventilation and by role modelling. Emotional-informational support can be considered a primary source – i.e., intimates - whereas club membership can be considered a secondary source of social support – i.e., knowledgeable others. This possibly explains the added value of both emotional-informational and club membership towards each other. Yet, it should be noted that club membership reflects more than the level of social support, as previous research also showed its link to socioeconomic status. For example, it was shown that adolescent girls with lower socioeconomic status were less likely to participate in a sports club [45]. Therefore, it is possible that a lack of club membership also reflects a lower socioeconomic status, which in turn negatively affects self-reported health [46]. In line, during the COVID-19 outbreak it was found that university workers living in smaller homes reported higher levels of anxiety-depression-and stress symptoms [47]. Also, for college students during the COVID-19 outbreak, economic disadvantage was associated with higher stress levels [48]. Perhaps, the higher levels of stress found in the present study for students without club membership are also partially explained by their socioeconomic status, but more research is needed.

A strength of the current study is that two measurements within two cohorts are included, enabling controls for baseline measurements and potential fluctuations throughout the academic year described in the literature [40]. Moreover, gender and study performance were controlled for, which are known to be correlated with stress levels [20, 22, 36]. Even though study performance was taken into account, the meaning of having obtained all possible credits up until the final measurement differed between cohorts. Due to the outbreak, exams were postponed which resulted in a higher number of exams that still had to be completed in the last part of the academic year. Therefore, the present study controlled for study performance of students regardless of the COVID-19 outbreak, in order to rule out potential effects of differences in performance level between students of different cohorts. However, this does not exclude the potential stress caused by postponed exams during the outbreak as described in a previous study [49]. Further, a limitation of the present study is the lower response-rate for cohort 2019. This was possibly due to the COVID-19 outbreak, which resulted in a fully online data collection instead of a combination of data collection in class and online. Even though we were able to control for student characteristics in the analyses, a bias may still be present due to this lower response-rate. The present study focused on Year-1 students, and this appears to be a relevant group since students in early stages of medical and dental school seem susceptible to the negative impact of the COVID-19 outbreak [19, 50]. Whether the results described in the present study will be similar for students in advanced stages of medical school needs to be further investigated. This also applies to the generalizability of the results to students from other schools, since the relation between social support and perceived stress may be different for medical students compared to students from other schools due to the higher prevalence of mental distress in medical students compared to their age-matched peers [1, 2].

Even though medical schools are not able to change the current COVID-19 crisis or any future crisis, they can help students get through it. Our findings suggest that in times of crisis medical students’ wellbeing can benefit from social support. With the COVID-19 pandemic still not being over and potential new lockdowns are possible, medical schools could play a more active role themselves by further exploring the (digital) options to provide different kinds of social support to its students. The first type of social support entails companionate care and instrumental assistance, which can be provided through one-on-one (online) mentoring with a faculty member [51], or a peer [52, 53]. The second type of social support has more to do with enabling ventilation with peers and role modelling, some sort of social embeddedness. Medical schools can achieve this by for example creating online communication platforms [54], or by stimulating peer relationships by promoting cooperation amongst students in the medical school program [55, 56]. All in all, when implementing (online) education, medical schools should not only focus on qualification but also on the social functions of education.

To conclude, the present study provides solid evidence of COVID-19 induced stress in medical students, especially among those with less social support. Findings of the present study are in line with an existing model describing the buffering effect of social support on crisis-induced stress and therefore go beyond the current COVID-19 pandemic. Medical schools can optimize social support for its students by offering social support on different levels in order to minimize the negative impact of future global, national or individual crises.

## Data Availability

For the sake of confidentiality, full data are not publicly accessible. Additional
anonymized data is available upon reasonable request to the authors.

## Acknowledgements

We thank Daphne Pol, who helped with data collection and data entry of cohort 2018 for our study. We also wish to thank David van Klaveren for helping us with our statistical analyses.

---

**S1 Table.**
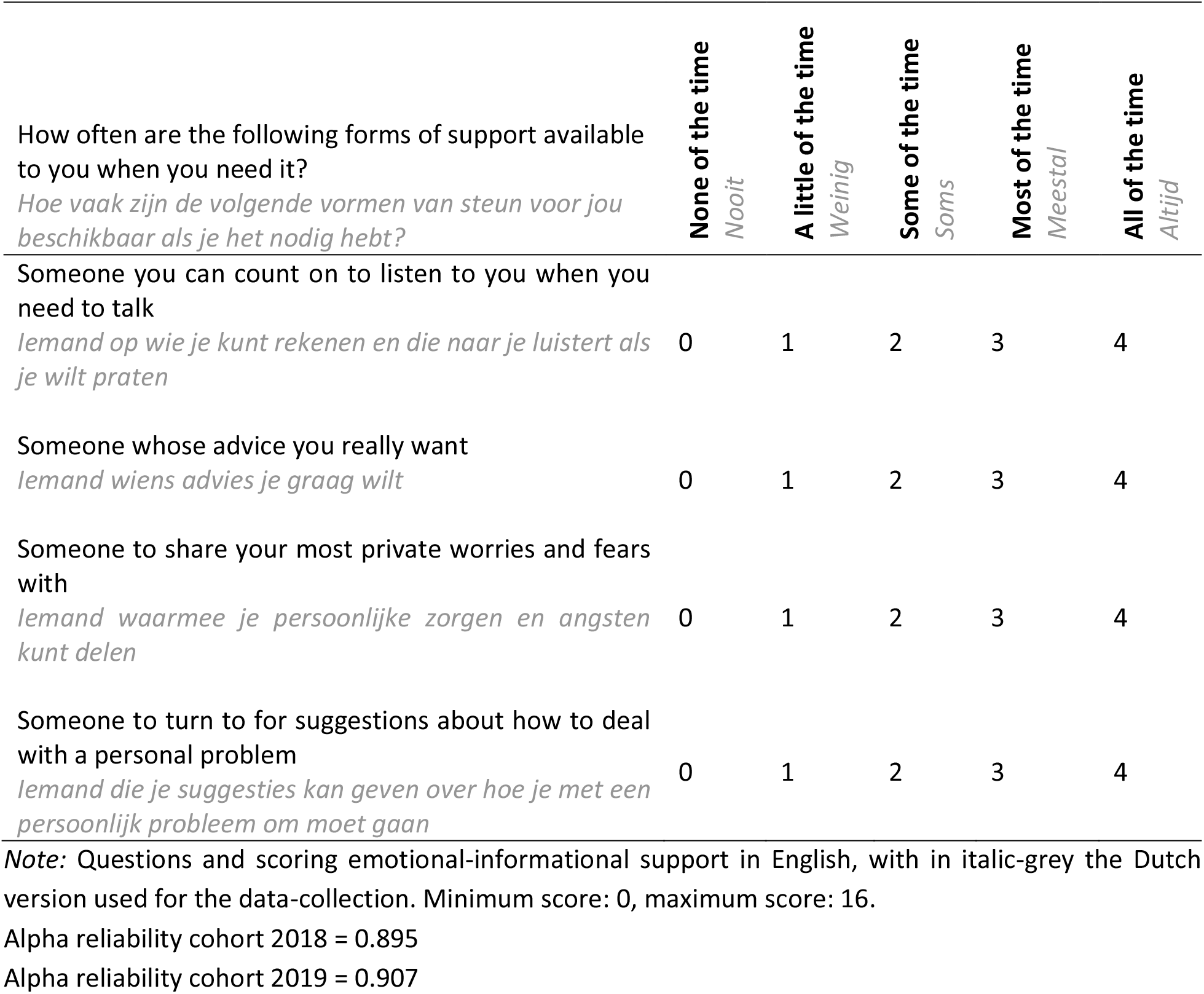
Measurement emotional-informational support.

